# Investigating adiposity in childhood and adulthood on later life sleep health: a lifecourse Mendelian randomization study

**DOI:** 10.64898/2026.08.27.26361310

**Authors:** Sweta Pathak, Tom G Richardson, Eleanor Sanderson, Nikhil Arora, Linn Beate Strand, Bjørn Olav Åsvold, Laxmi Bhatta, Ben Brumpton

**Affiliations:** HUNT Center for Molecular and Clinical Epidemiology, Department of Public Health and Nursing, Norwegian University of Science and Technology, Trondheim, Norway; MRC Integrative Epidemiology Unit, Population Health Sciences, Bristol Medical School, University of Bristol, United Kingdom; Department of Endocrinology, Clinic of Medicine, St. Olavs Hospital, Trondheim University Hospital, Trondheim 7030, Norway; HUNT Research Centre, Department of Public Health and Nursing, NTNU Norwegian University of Science and Technology, Levanger, Norway; Clinic of Medicine, St. Olavs Hospital, Trondheim University Hospital, Trondheim, Norway; Department of Public Health and Nursing, NTNU Norwegian University of Science and Technology, Trondheim, Norway

**Keywords:** childhood, adulthood, obesity, insomnia, morning chronotype, medelian randomization

## Abstract

**Background:** Higher Body Mass Index (BMI) is an established risk factor of sleep disturbance. It is not known if the effect is homogeneous across the lifecourse or if there is a particular time point in life that might be best to target.

**Methods:** Two-sample Mendelian randomization (MR) was used to investigated the effect of childhood adiposity (adjusting on adulthood adiposity and obstructive sleep apnea (OSA)) on insomnia, morning chronotype, sleep duration, daytime sleepiness and daytime napping. Similarly, total, and direct effect of adulthood adiposity on these outcomes was explored. We used summary statistics from a genome-wide association study (GWAS) of UK Biobank for childhood and adulthood adiposity (n=453,169) and large-scale consortia of OSA (Million Veteran Program) (n=410,268), insomnia, and chronotype (23andMe) (n=1,978,022 and n=248,1000, respectively).

**Results:** Two-sample univariable MR analysis provided no evidence of an effect of genetically predicted childhood adiposity on later life insomnia (Odds ratio (OR)= 0.94, 95% Confidence interval (CI)= 0.87, 1.03). Whereas, multivariable MR (adjusted for adulthood adiposity) analysis provide strong evidence of direct protective effect of genetically predicted childhood adiposity on later life insomnia (OR= 0.70, CI= 0.64, 0.77). Further, both in univariable and multivariable MR, a strong positive effect of increased childhood body size on morning chronotype was observed (OR= 1.16, CI= 1.01, 1.33 and OR= 1.36, CI= 1.15, 1.62, respectively) after accounting for adulthood body size. In both analysis the estimate did not change considerably after aditionally adjusting for OSA. However, childhood and adulthood adiposity found to be associated with OSA and OSA with insomnia. In both univariable and multivariable analysis, increased body size in adulthood increased the risk of having insomnia and a morning chronotype.

**Conclusions:** The findings suggest that higher body size in childhood is not a risk factor for later life insomnia, whereas higher body size in adulthood was. Further, if healthy body size is maintained in adulthood, high childhood adiposity may decrease the risk of insomnia and increase the risk of being a morning person in later life.

## BACKGROUND

Good sleep quality is basic human need for health and well-being ^1–3^. Studies suggest that sleep disorder are associated with neural development, memory, cardiovascular, metabolic function, cancer, and premature mortality ^4–7^. Sleep traits such as insomnia, chronotype (morningness or eveningness), sleep duration, daytime sleepiness explain individual sleep health among which insomnia is most common type of sleep disorder. Insomnia includes difficulties to initiate and maintain sleep or poor quality of sleep ^8,9^. It is said that 30-40% of the world’s adult population is affected by insomnia ^10,11^. This growing evidence of sleep abnormality makes it an emerging global epidemic. Increasing body mass index (BMI) is an established risk factor of sleep disturbance ^12^ therefore it is important to look at adiposity at different time points in the lifecourse to understand if the effect is homogeneous or if there is a particular time point in life that might be best to target. However, no studies have investigated the direct effect of adiposity at childhood and adulthood in multivariable MR designs. This is important because studing the total effects of childhood or adulthood adiposity alone without controlling for the other, can obscured the true causal effect of these individual time points. If adiposity has different effects at different time points in the life course, prevention stratergies which target adiposity to improve sleep health should adapt.

Obstructive sleep apnea (OSA) is a condition were breathing repeatedly stops and starts during sleep and it is considered to be one of the key mechanisms from obesity to insomnia^13^. Several studies have suggested that there is a causal relationship between adiposity with OSA ^14–16^. Similary, studies shows that OSA and insomnia are often co-morbidities, with 39- 50% of patients with OSA having insomnia ^17–20^. Therefore, in order to investiagate the direct causal effect between adiposity and sleep disorder, it is important to consider OSA.

In this study, we used multivariable Mendelian Randomization (MR) in a lifecourse approach, which uses genetic variants (single nucleotide polymorphisms; SNPs) as instrumental variables (IV) for specific time-points (Figure 1), to investigate the direct effect of childhood adiposity taking account of adulthood adiposity and additionally OSA on later life sleep health (insomnia, morning chronotype/morningness, sleep duration, daytime sleepiness, daytime napping). This approach may also reduce residual confounding and reverse causation which is the common source of bias in observational studies. ^21–23^

**Figure 1.**
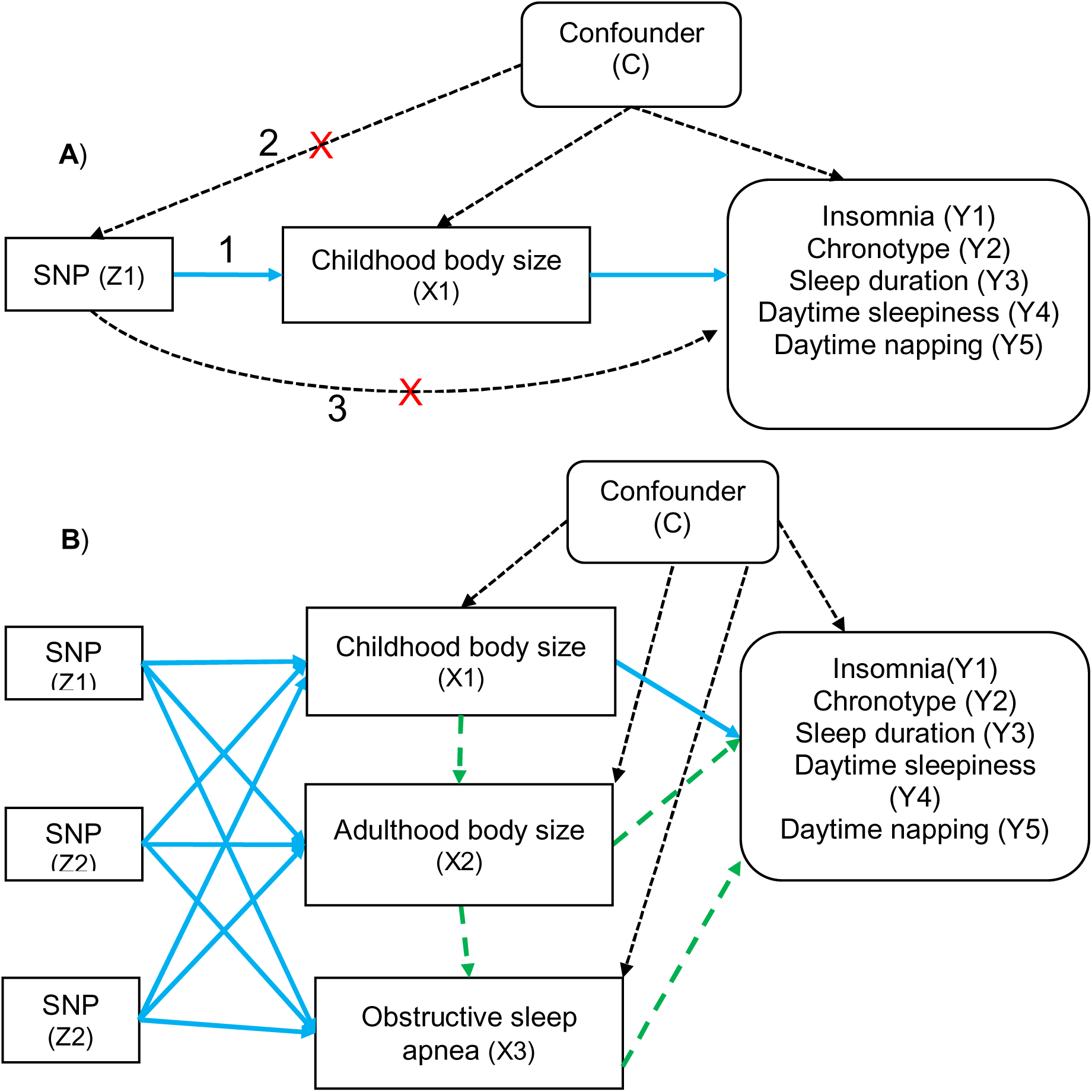
legend. A directed acyclic graph of the two-sample MR framework. **A)** Univariable MR framework in the relationship (total effect) of childhood adiposity (X1) on later life insomnia (Y1), chronotype (Y2), sleep duration (Y3), daytime sleepiness (Y4), and daytime napping (Y5). The three core MR assumptions are 1) the relevance - the SNP (Z1) must be associated with the exposure (X1), 2) the independence - the SNP (Z1) must not be associated with confounders of the exposure (X1) - outcome (Y1, Y2, Y3, Y4, and Y5) relationship, and (3) the exclusion-restriction - the SNP (Z1) must only be associated with the outcome (Y1, Y2, Y3, Y4, and Y5) via the exposure (X1) **B)** Multivariable MR framework where the direct effect pathway is the solid line from childhood adiposity on later life insomnia, chronotype, sleep duration, daytime sleepiness, and daytime napping, while the total effect pathway includes both the solid line from childhood and the dotted line from adulthood adiposity and obstructive sleep apnea on later life insomnia, chronotype, sleep duration, daytime sleepiness, and daytime napping. Z1 - genetic variants associated with the first exposure of interest (X1), Z2 - genetic variants associated with the second exposure (X2), Z3 - genetic variants associated with the third exposure (X3).

## METHODS

### Study design

We used two-sample univariable and multivariable MR approach to quantify the total and direct effect of childhood adiposity on later life sleep health (insomnia and morning chronotype) controlling for adulthood adiposity and OSA ^24,25^. Additionally we investigated the effect of childhood adiposity (adjusting on adulthood adiposity and OSA) on sleep duration, daytime sleepiness and daytime napping. Similarly, the total, and direct effect adjusting for childhood adiposity and OSA of adulthood adiposity on these outcomes are explored (Figure 1). We have presented a flow chart to explain the workflow of the two-sample MR design in Figure 2.

**Figure 2.**
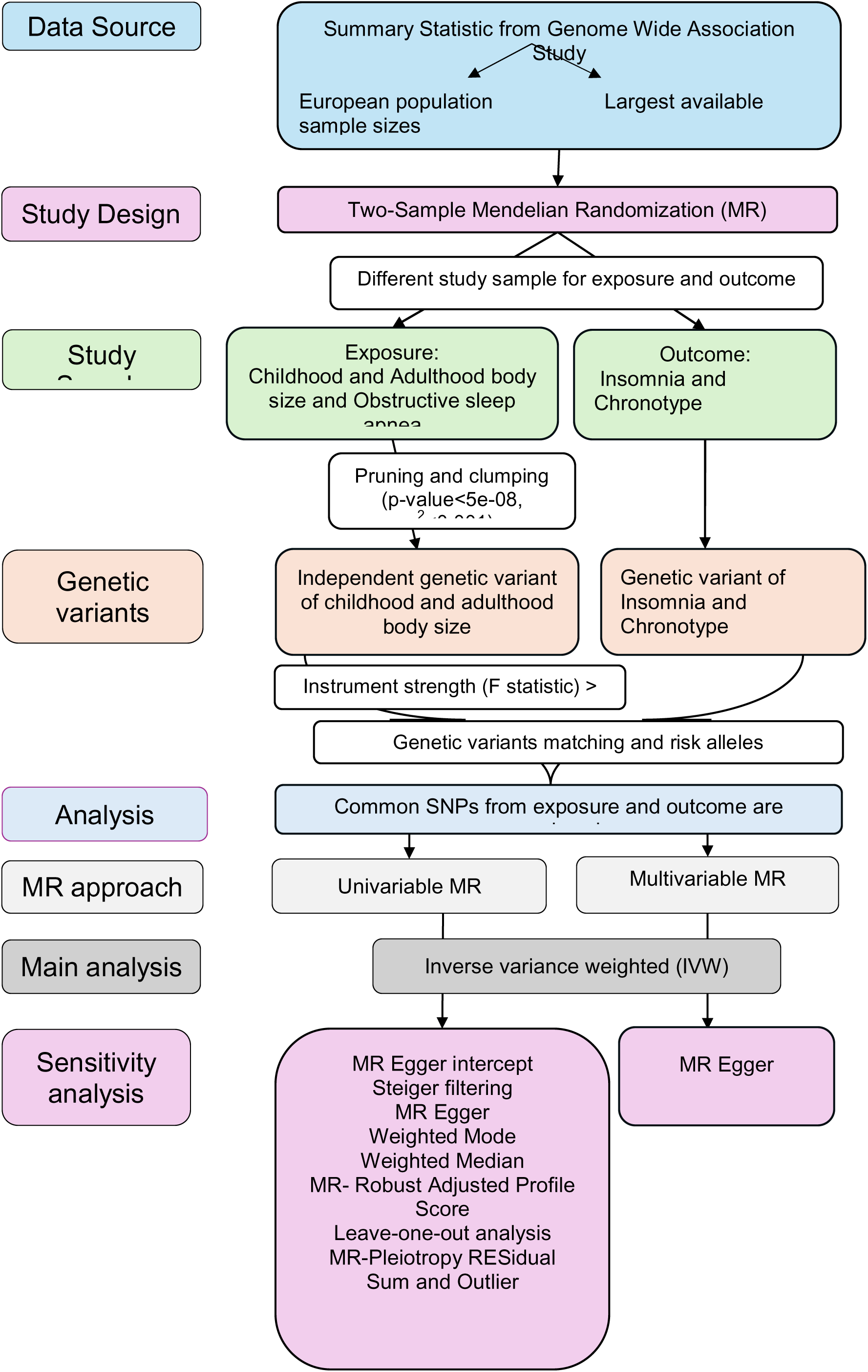
Flow chart showing potential mechanism of two-sample MR analysis.

### Data resources

To perform two-sample MR, we used summary-level data from large-scale genome-wide association studies (GWASs) for the exposure and the outcomes, conducted on non-overlapping cohorts and in same underlying population (European) ^26^. An overview of the data used is provided below (Table 1).

**Table 1.**
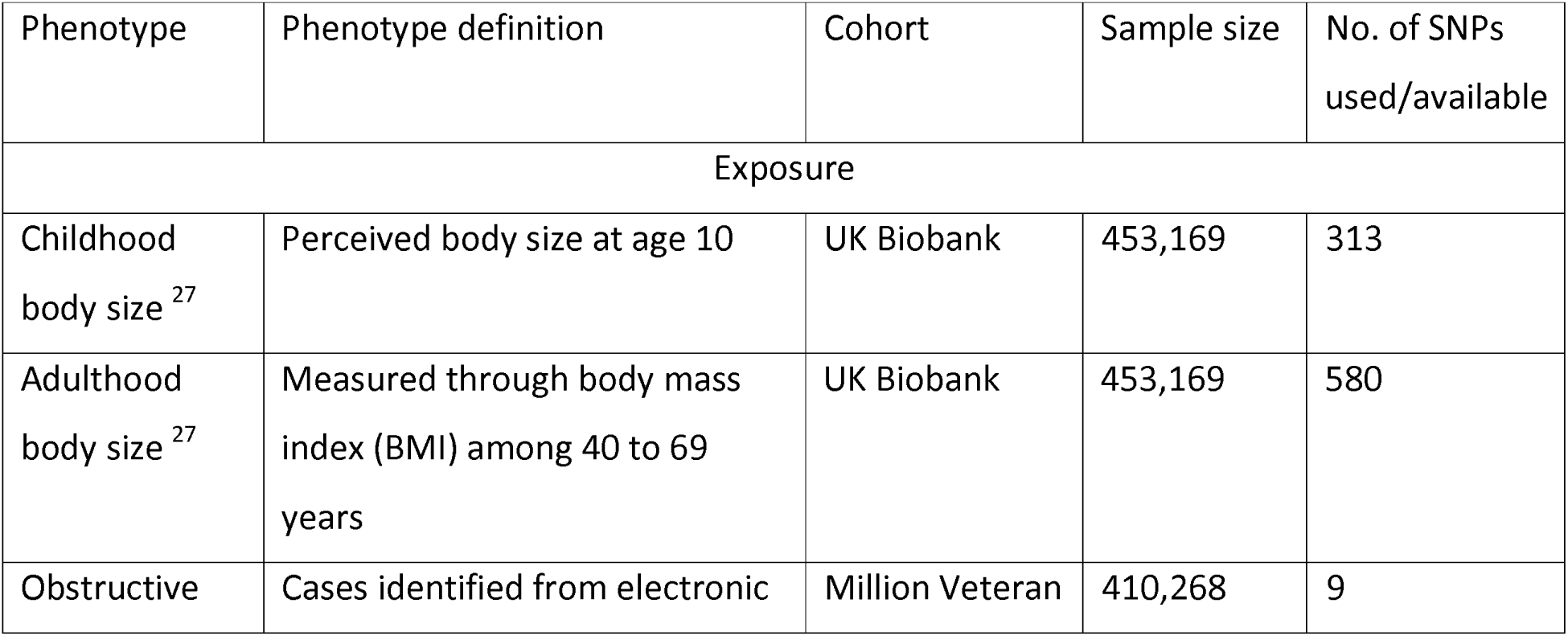
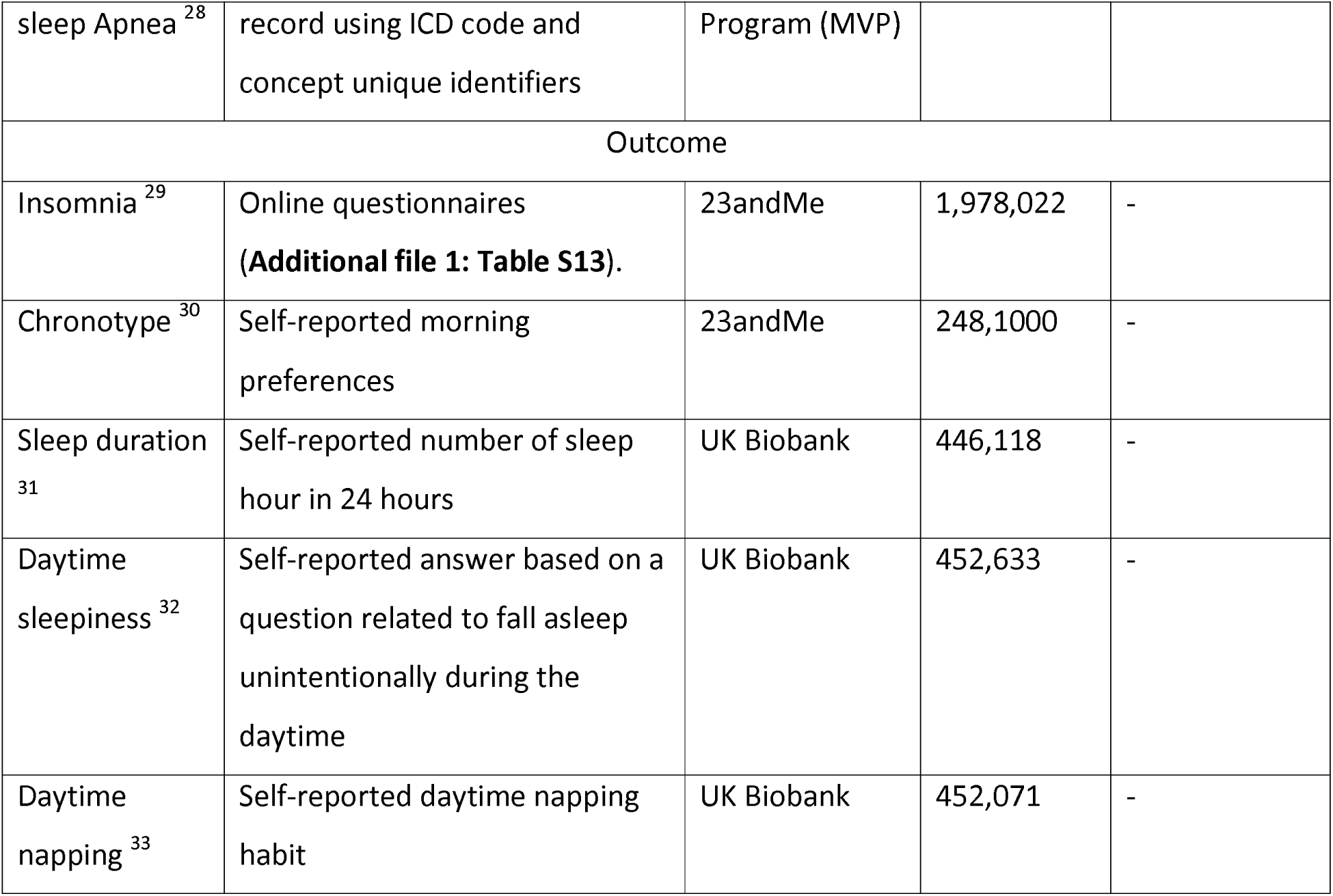
Large-scale GWAS used for adiposity and sleep traits/disorders.

**Table 2.**
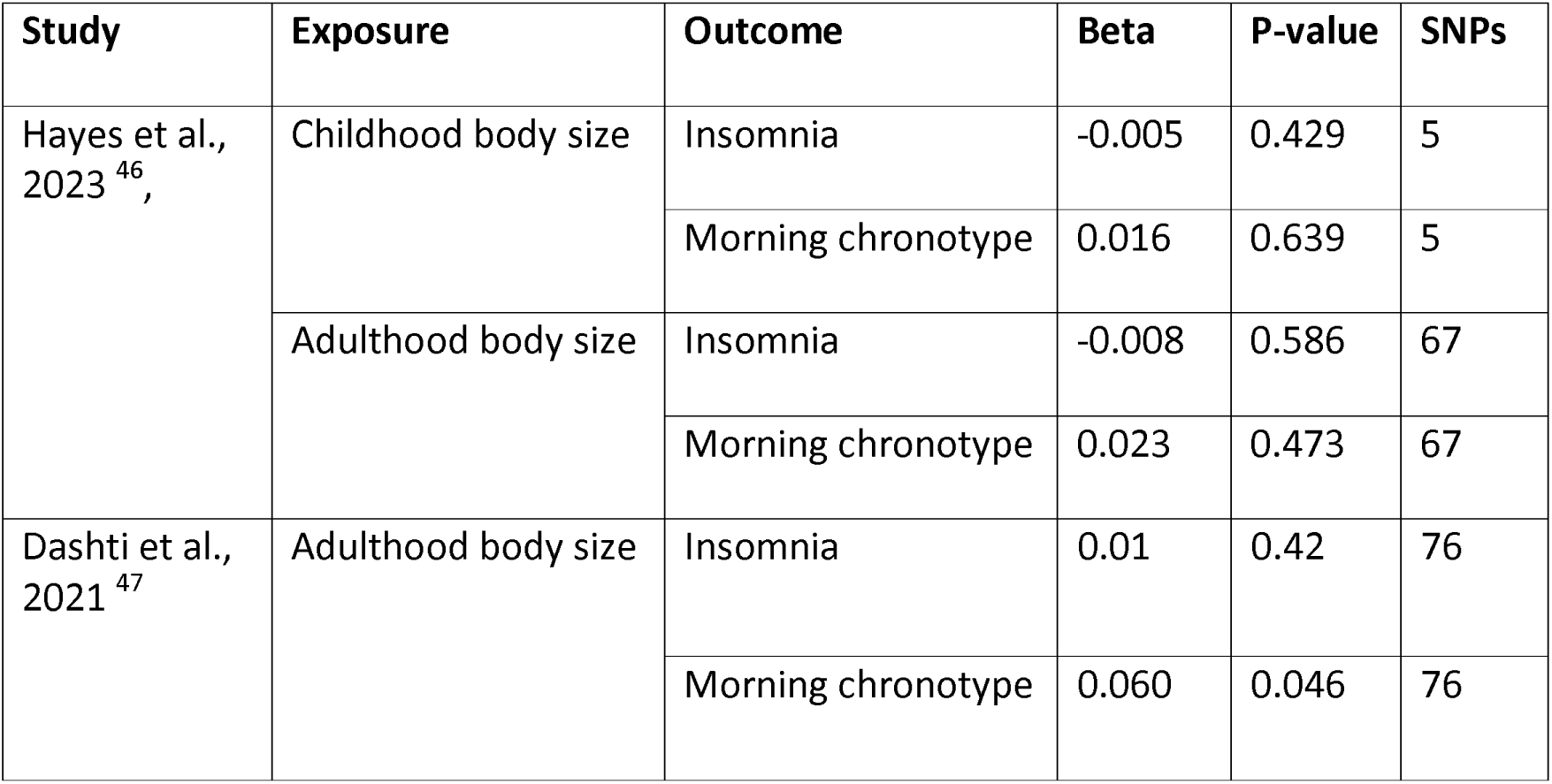
Summary of previously published two-sample univariable MR studies for adiposity and sleep traits.

### Genetic instruments for childhood and adulthood body size

Independent genetic variants (SNPs) of childhood and adulthood body size were identified from GWAS performed on 453,169 European ancestry in the UK biobank study, adjusted for age, sex, genotyping chip, relatedness and population stratification ^27^. To identify indepentend SNPs associated with childhood and adulthood body size, the results were clumped using the 1000 genomes reference panel of phase 3 (version 5) and the criteria r^2^ <0.001 and p-value <5x10^-08^ in PLINK. Finally, 313 and 580 independent SNPs were identified as genetic instrument for childhood and adult body size, respectively ^27,34^.

Adult body size between the aged of 40 and 69 years was estimated using body mass index (BMI), which is calculated by dividing weight (to the nearest 0.1 kilogram) by height (measured in whole centimetres) squared. At the same time people were asked to describe their childhood body size when they were 10 year old as thinner, plumper, and about average, which described childhood body size ^27^. Finally, participants who have both information of childhood and adulthood body size were taken for further analysis ^27^. To make both childhood and adulthood body size comparable, adult BMI was also categorized as thinner, plumber, and about average on the same proportions as childhood body size variables ^27^. Among 10,000 randomly selected unrelated participants in UK Biobank a genetic correlation matrix was derived and used to calculate the genetic correlation between childhood and adulthood body size (rg=0.61) ^27^, indicating substaintial genetic differences between.

To account for the limitation of using recalled childhood body size, validation analysis was carried out in Avon Longitudinal Study of Parents and Children (ALSPAC) ^27^, the Young Finns study ^35^ and the Trøndelag Health (HUNT) study ^36^, where only people with early life and later life measures were included ^27^. Validation analysis showed that, the genetic instrument for childhood body size from UK Biobank was found to be strong predictor of childhood BMI and less predictive of adulthood BMI with a genetic correlation (rg) of 0.61 ^27,35,36^.

### Genetic instruments for Obstructive Sleep Apnea

Genetic variants associated with OSA was retrieved from GWAS performed in the Million Veteran Program (MVP) among 410,268 European participants aged 64 years old on average^28^. Cases of OSA were identified from Veterans Affairs electronic health record by using multimodal automated phenotyping procedure (predictive probability of phenotype ^37^), using International Classification of Disease (ICD) -9 code 327.23 and ICD-10 code G47.33, and concept unique identifiers C0520679 ^28^.

To identify independent genome-wide significant ( p-value <5x10^-08^ ) SNPs, MVP GWAS used the clumping criteria of r^2^ = 0.1. In our study, to make similar clumping criteria in all exposure we then again applied clumping criteria of r^2^ <0.001 ^28^. From the MVP GWAS, 12 independent significant SNPs were identified and after applying strict clumping criteria of r^2^ <0.001, 3 genetic variants out of 12 were removed due to Linkage disequilibrium (LD) with other variants or absence from LD reference panel and among 3 removed variants 2 were multi-allelic snps not in LD, so, they were again added. Finally 11 independent genetic variant were used as an instrumental variable of OSA ^28^.

### Genetic instruments for sleep traits

We retrieved summary statistics for insomnia, morning chronotype, sleep duration, daytime napping, daytime sleepiness from large scale GWAS of the European population.

Summary statistics for insomnia were obtained from GWAS performed in the 23andMe cohort among 1,978,022 (484,176Lcases and 1,493,846 controls) unrelated European participants of age 20 to 80. Logistic regression was performed adjusting for age, sex, genotype array and the first five genetic ancestry principal components (PCs) ^29^. The GWAS of insomnia had a λ =1.00, LD intercept of 1.15 and the SNP-heritability (h^2^SNP) of insomnia was estimated to be 8.15% within the dataset ^29^. In this study, cases of insomnia was defined from participants answers to multiple questionnaires based on an online survey. Seven questions related to insomnia were asked and participants with positive answers in any of the seven questions were consider as cases and participants who did not provide either positive or uncertain answer were additionally asked 5 more questions to be consider as control ^29^ (Additional file 1: Table S13).

Summary statistic of chronotype were obtained from a GWAS performed in the 23andMe cohort among 248,100 (120,478 cases and 127,622 controls) unrelated participants of European-ancestry aged 20 to 80 ^30^. Phenotypic information on chronotype (mornig person or night person) is self-reported based on the question “Are you naturally a night person or a morning person?” with the possible answer, first - “Night owl”, “Early bird” and “Neither” and second - “Night person”, “Morning person”, “Neither”, “It depends” and “I’m not sure”. ^30^. Summary statistic of chronotype was generated using logistic regression analysis (additive model) adjusting for age, sex, the first four principal component, and a categorical variable representing genotyping platform. ^30^.

The GWAS of sleep duration was performed in UK biobank among 446,118 participants of european ancestry ^31^. Sleep duration was self-reported by participants based on a question related to number of hours sleeping (including napping) in a day (24 hours). GWAS of sleep duration had the SNP-heritability (h^2^SNP) of 9.8% within the dataset. Sleep duration was assessed on a continuous scale based on 3 categorizes; short (6⍰h or less), normal (7 or 8⍰h), or long (9⍰h or more) sleep duration ^31^. The GWAS of sleep duration was performed using a linear mixed model adjusted for age, sex, 10 principal components of ancestry, genotyping array, and genetic correlation matrix with a maximum per SNP missingness of 10% and per sample missingness of 40% ^31^.

We retrieved SNP-daytime sleepiness associations from GWAS performed in 452,071 European participants from the UK Biobank ^32^. Daytime sleepiness is self-reported based on a question on falling asleep unintentionally during the daytime. In the analysis it was treated on a continuous scale based on four categorizes (“Never/rarely”, “sometime”, “often”, and “all of the time”)^32^. GWAS of self-reported daytime sleepiness was performed using linear mixed regression model adjusted for age, sex, genotyping array, ten principal components (PCs) of ancestry ^32^. The SNP-heritability of self-reported daytime sleepiness was reported to be 6.9% ^32^.

Summary statistic on day-time napping was extracted from GWAS performed in UK Biobank among 452,633 European participants ^33^. Information on day time napping was self-reported from the question “Do you have a nap during the day?”, where the participants answered never/rarely, sometimes, and always. In the analysis the variable was treated on a continuous scale . The GWAS was performed using a linear mixed model adjusted for age, sex, 10 principal components of ancestry, genotyping array, and genetic correlation matrix with a maximum per SNP missingness of 10% and per sample missingness of 40% ^33^. The GWAS of day-time napping had a LD score regression intercept of 1.04 and SNP-heritability of 11.9% ^33^.

## STATISTICAL ANALYSIS

### Testing the causal association of adiposity on insomnia and chronotype

Two sample-Mendelian Randomization (MR) analysis was used to explore the causal pathway between adiposity and insomnia and chronotype by using genetic variants (SNPs) of adiposity as instrumental variables (IV).

Univariable MR was used to estimate the total effect of genetically predicted childhood body size on insomnia and morning chronotype which is presented as model 1. While Multivariable MR was used to estimate the direct effect of childhood adiposity on insomnia and morning chronotype while adjusting for adulthood adiposity is model 2. Additionally, Model 3 (multivariable MR) was built to assess the effect of childhood adiposity on insomnia and morning chronotype while adjusting for OSA in addition to adulthood adiposity. As OSA has been associated with both insomnia ^17–19^ and adiposity ^14–16^, we wanted to control for its effect to estimate the direct effect of childhood adiposity on insomnia. Similarly, total, and direct effect of adulthood adiposity adjusting on childhood adiposity and OSA on insomnia and morning chronotype were also estimated.

Inverse variance weighted (IVW) method was used as a primary analysis to estimate the causal association of the exposure on outcome both in univariable and multivariable MR. Each genetic variant provides an independent ratio estimate of the association which is calculated by the wald ratio (SNP-outcome association divided by SNP-exposure association), then these ratio estimates are combine together in a fixed effect meta-analysis which give the IVW estimate ^38^. To check the compatibility of each ratio estimate including in the IVW estimate, we also performed a heterogeneity test and presented the Q-statistic, which has a chi-square distribution with n-1 degrees of freedom. Before summary-level analysis, effect alleles of childhood body size were matched with adulthood body size, OSA, insomnia and morning chronotype to avoid combining incorrect directions of effects calculated by Wald ratio for each SNPs, which would later lead to bias in the estimate. If all the genetic variants used in MR are valid , the IVW estimate is most powerful ^39^. Valid instruments can provide independent and unbiased causal estimates ^40^. We checked the strength (relevance assumption) of the genetic instrument by calculating conditional and unconditional F-statistic of childhood and adulthood adiposity ^40^ . The F-statistics was >10 suggesting that weak instruments were unlikely to substantially bias the analysis (Additional file 1: Table S7).

To quantify the robustness of the IVW estimate, we applied sensitivity analysis to understand if our causal effect estimates are biased because of pleiotropy (which is the effect of genetic variants on the outcome through different phenotypes other than exposure of interest). As a sensitivity analysis, MR egger estimates were calculated, and additionally for the univariable MR, weighted mode and weighted median, leave-one-out analyses, MR-Pleiotropy RESidual Sum and Outlier (MR-PRESSO) ^41^, MR-Robust Adjusted

Profile Score (MR-RAPS) ^42^, and Steiger filtering was performed to explore potential horizontal pleiotropy. To explore potential horizontal pleiotropy MR Egger intercepts was also calculated.

Data were analysed using the ‘MendelianRandomizatioń package in R statistical software, and all the IVW causal estimates were presented in forestplots using the ‘forestplot’ package in R (Figure 3).

**Figure 3.**
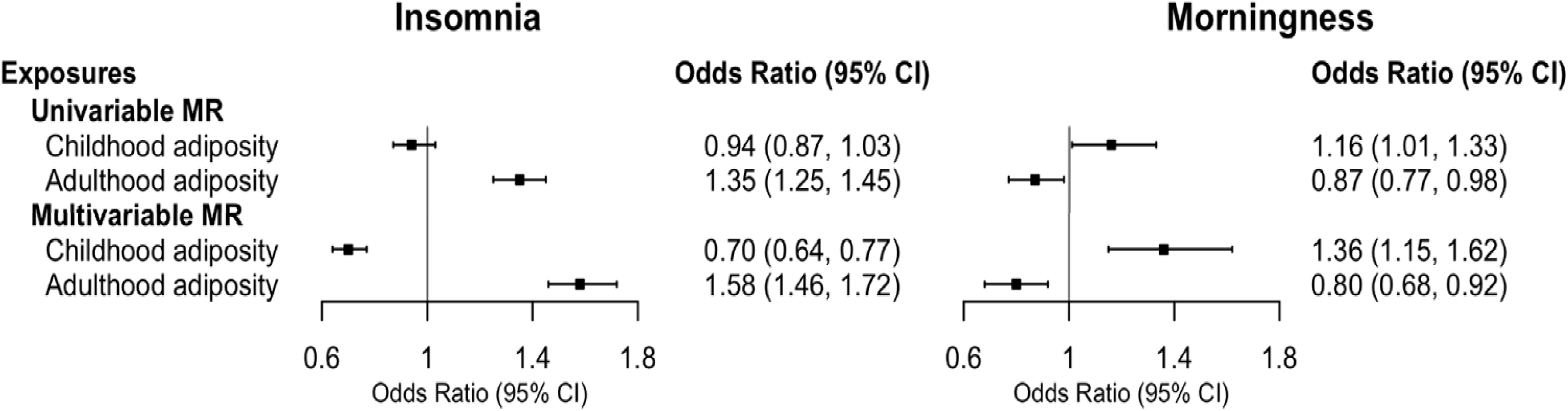
legend. Forest plot illustrating the total and direct causal estimates of childhood adiposity and adulthood adiposity on insomnia and morning chronotype. The univariable MR represents the total causal estimate and multivariable MR represents direct causal estimates after adjusting for adulthood adiposity. The causal estimates are presented as odds ratio with 95% CI. Abbreviations: MR, Mendelian randomization; CI, confidence interval

### Testing the causal association of adiposity on sleep related traits

We applied the same analysis procedure above to estimate the total and direct effect of childhood and adulthood adiposity on sleep duration, daytime sleepiness, and daytime napping. Two sample-MR generally use exposure- and outcome-associations from non-overlapping participants/cohort to avoid potential biases from overfitting ^43,44^. Here, our exposure; childhood and adulthood adiposity and outcome; sleep duration, daytime sleepiness, and daytime napping both are from UK biobank study, therefore we assume that the estimate might be biased from overfitting (Additional file 1: Table S12). Therefore, we presented this result as an additional analysis which required further validation conducted on non-overlapping samples.

## RESULTS

Total effects of childhood and adulthood body size on insomnia and morning chronotype Two-sample univariable MR analysis provided no evidence of an effect of genetically predicted childhood adiposity on later life insomnia (Odds ratio (OR)= 0.94 per change in body size, 95% Confidence interval (CI)= 0.87, 1.03). However, higher body size in adulthood was associated with an increase in insomnia (OR= 1.35 per change in body size, 95% CI= 1.25, 1.45). Whereas, estimates suggest a positive effect between genetically predicted childhood adiposity and later life morning chronotype (OR= 1.16 per change in body size, 95% CI= 1.01, 1.33) and negative effect between adulthood adiposity and morning chronotype (OR= 0.87 per change in body size, 95% CI= 0.77, 0.98) (Additional file 1: Table S1).

### Direct effects of childhood and adulthood body size on insomnia and morning chronotype

Two-sample multivariable MR analysis provide strong evidence of direct protective effect of genetically predicted childhood adiposity on later life insomnia after adjusting for adulthood body size (OR= 0.70 per change in body size, 95% CI= 0.64, 0.77). Similar to the univariable MR result, higher body size in adulthood (accounting for childhood body size) increased the risk of having insomnia (OR= 1.58 per change in body size, 95% CI= 1.46, 1.72). Moreover, we observed a stronger positive effect of higher childhood body size (accounting for adulthood body size) on morning chronotype (OR= 1.36 per change in body size, 95% CI= 1.15, 1.62) and negative effect of adulthood body size (accounting for childhood body size) on morning chronotype (OR= 0.80 per change in body size, 95% CI= 0.68, 0.92), which is consistent with univariable MR result (Additional file 1: Table S1).

In additional analysis (model 3), were in addition to childhood/adulthood body size also adjusted for OSA to see direct effect of adiposity on sleep, the estimate did not change considerably (Additional file 1: Table S1). Before, constructing model 3 ,the causal direction between OSA and insomnia was also tested using two-step univariable MR, two-step multivariable MR and a directionality test ‘Steiger method’ ^45^ which tests that the variance explained of OSA is greater than insomnia which would suggest that the causal direction from OSA to insomnia is true ^45^ (Additional file 1: Table S11). In the first step of the two-step univariable MR, both childhood and adulthood adiopoisity was associated with OSA (OR= 1.73, 95% CI= 1.55, 1.92 and OR= 2.31, 95% CI= 2.13, 2.50 , respectively) (Additional file 1: Table S10). However in the multivariable MR, only adult adiposity was associated (OR= 2.19, 95% CI= 1.96, 2.43) (Additional file 1: Table S10). In the second step of the two-step univariable MR, OSA was associated with insomnia (OR= 1.61, 95% CI= 1.44, 1.81).

However, in the multivariable MR there was no clear association (Additional file 1: Table S9).

### Sensitivity analysis

To test the robustness of the IVW results in the univariable and multivariable MR analysis, we additionally performed sensitivity analysis. In univariable MR, MR Egger, Weighted Median, and Weighted Mode, and MR-RAPS estimates slightly vary from univariable IVW estimates (Additional file 1: Table S2, S4 and S5). In leave-one-out analysis, where the causal effects of exposure on the outcome was calculated by removing each SNPs from the analysis, the results did not shows any strong effects of any particular SNP (Additional file 1: Figure S1). We further tested for outlier using MR-PRESSO analysis and no outlying SNPs were detected. In multivariable MR, MR egger estimates (Additional file 1: Table S2) were similar to the finding of multivariable IVW estimates. The MR egger intercept results (Additional file 1: Table S3) did not indicate any strong horizontal pleiotropy. However, evidence of heterogeneity was observed across all analyse (Additional file 1: Table S8).

### Additonal analysis

In two-sample univariable MR analysis, a positive association of genetically predicted childhood and adulthood adiposity on later life daytime sleepiness was observed. However, higher body size during childhood tended to reduce sleeping hours and the habit of daytime napping. Whereas, a higher adult body size reduced sleeping hours but increased the habit of daytime napping (Additional file 1: Table S12).

In two-sample multivariable MR analysis, there was no evidence of a direct effect of childhood adiposity (accounting for adulthood adiposity) on sleep duration and daytime sleepiness. However, we observed a protective effect on daytime napping. Similar to the finding of univariable MR, higher body size during adulthood (accounting for childhood adiposity) lowered sleeping hours and increased daytime sleepiness and daytime napping (Additional file 1: Table S12).

## DISCUSSION

In this lifecourse MR study, we examined the total and direct effect of childhood and adulthood adiposity on insomnia and morning chronotype. We found evidence suggesting that genetically predictive childhood adiposity has direct (adjusting for adulthood adiposity) protective effect on insomnia. However, our estimates still show that high adulthood adiposity is a strong risk factor for insomnia, with or without adjusting for childhood adiposity. Additionally, despite observing strong associations in two-step univariable MR between childhood and adulthood adiposity and OSA, and OSA and insomnia, we did not observe that controlling for OSA materially changed our main findings. Further, we observed that higher body size during childhood increased the chance of being a morning person in later life whereas higher body size during adulthood decreases morning preference.

To explore the effect of adiposity across the lifecourse and to understand if the effect is homogeneous or if there is particular time point in life that has effect on later life sleep. It is important to disentangle the association between childhood and adulthood adiposity with sleep. We applied multivariable MR, which helps to estimate the direct effect of adiposity at two different timepoints. Testing the effect of adiposity at two different time points (childhood and adulthood), may pin point which time point could be critical for life course public health interventions.

The aim of our study was to understand how childhood obesity affects later life sleep health while addressing the influence of adulthood obesity. To our knowledge, except our study no other genetic study has explore direct effect of childhood/adulthood adiposity on insomnia and morning chronotype. Therefore an exact comparison of the finding of our study is not possible. However, two previously performed genetic studies have explored univariable effects of childhood and adulthood adiposity on sleep trait ^46,47^, where Hayes et al. ^46^ found no evidence for an effect of genetically predicted childhood adiposity on insomnia which is similar to the finding of our univariable MR result. However, after accounting for adulthood adiposity, a strong protective effect of childhood adiposity on later life insomnia was observed in our study. Similar protective effect of childhood adiposity has been observed in some other phenotype including mental health ^48^. Hayes et al. ^46^ and Dashti et al. ^47^ has found no any association between adulthood body size and insomnia but in contrast with this finding, our study found that higher adult body size increases risk of insomnia. Furthermore, Hayes et al. found no clear effect of childhood and adulthood adiposity on morning chronotype which is inconsistent with our finding.

A possible reason for the differences in results between our study and previously performed studies might be due to the differences in study design, where we used a multivariable MR approach, as studing the total effects of childhood or adulthood adiposity alone without controlling for the other, can obscured the true causal effect of these individual time points. Studies have found that children who are obese during childhood are more likely to be obese during adulthood ^49^. Also, a genetic study has reported that childhood and adulthood adiposity are 61% corelated ^27^. Therefore it was important to do multivariable MR to see direct effect of childhood and adulthood adiposity on later life sleep health.

Furthermore, some studies have highlighted the association of OSA and insomnia ^17–19^ so, inorder to acount for OSA we included OSA in addition to childhood/adulthood adiposity (model 3) and our estimates did not change considerably. In our univariable two-step MR analysis, our estimates suggest that 1) higher adult body size is a risk factor for OSA, and 2) OSA was associated with insomnia, which is similar to the finding of several other observational study ^50–52^. However, in our multivariable two-step MR analysis after adjusting for childhood and adulthood body size no association between OSA and insomnia was observed. These analyses suggest that effect of OSA on sleep health is via adiposity.

Beside the above named genetic studies, no observational study has investigated the longitudinal effect of adiposity on insomnia and chronotype. However, in contrast to our study, a meta-analysis of observational cross-sectional studies conducted by Chan et al. ^53^, observed no association between body mass index and insomnia (OR = 1.07, p = .40). Additionaly, in another meta-analysis of randomized controlled studies and observational studies by Zhang et al. ^54^ the authors found that compared to people with a morning chronotype, people with a evening chronotype people had a larger body size, which is consistent with our finding.

The mechanism of effect from adulthood adiposity on insomnia and morning chronotype might be because of the phenomena where excess fat narrow the airway causing breathing difficulties ^55^ which can cause disturbance in sleep at night and make it difficult for people to wakeup in the morning. Also, people with higher body size are at higher risk of having co- morbidities, such as Gastroesophageal Reflux Disease (in which stomach acid flows back into the esophagus) ^56^ and depression and anxiety ^48^, which might effect people’s sleep health.

Result from addition analysis of sleeprelated traits indicate that childhood adiposity did not effect sleep duration and daytime sleepiness, however did indicate that high childhood adiposity increased daytime napping, which is similar to the finding from Hayes et al. study. Furthermore, Dashti et al. and Hayes et al. found a robust causal effect of higher adult BMI on increasing daytime sleepiness ^47,57^ which is similar to the finding of our study. However, estimates from our study suggest that higher body size during adulthood decreases sleeping hours and increased daytime napping.

### Strength and Limitation of this study

The key strength of our investigation is the use of two-sample lifecoursr MR framework, this is a unique feature of this study which enabled us to explore the independent lifecourse effects of each exposure on a single outcome ^25,58^. This is also a unique feature of this study which allows us to estimate the effect of same exposure at two different time points, which may help guide lifecourse health intervention. Furthermore the multivariable MR framework enable us to use large sample sizes from available GWAS, and also GWAS summary statistic from non-overlapping cohort for the exposure and outcome which help to minimize the potential bias due to overfitting ^44^. Additionally, in this design we use genetic variant as an instrumental variable for the exposure to help mitigate confounding and reverse causation. Furthermore, due to the large sample sizes and availbalitly of data we were able to thoroughly test MR assumptions and disentangle potentially strong confounders or mediators such as OSA.

This study, however, also has certain limitations. Childhood body size is perceived body size rather than a measured variable, where adult participants recalled their childhood body size. This may result in information bias and childhood body size may reflect different features of adiposity than adulthood adiposity. However, three validation studies were performed in order to account for the limitation of using perceived body size ^27,35,36^ which showed that the genetic instrument for childhood body size from UK Biobank was found to be a strong predictor of childhood BMI compared to the genetic instrument for adulthood BMI. Also, bias by population stratification, dynastic effects, and assortative mating may bias our results ^59^. To address population stratification, GWASs study (used as exposure and outcome in our study) has adjusted for PCs. We have shown that these effects on BMI are relatively small ^60^, however we have not explored these potential biases with insomnia or sleep related traits. Compared to national representative data sources, UK biobank participants are more likely to be older, female which might have introduced selection bias in our study ^61^. Finally, all the GWAS used in this study are based on European population which might limits the generalizability of results.

## CONCLUSION

In this lifecourse mendelian randomization study, childhood adiposity was not found as a risk factor for later life insomnia and the effect seems to be via adult adiposity. Further, our study suggest that childhood adiposity increases the chance of being a morning person in later life, if healthy body size is maintained during adulthood. Additionally, adult adiposity was found as a strong risk factors for OSA and suggest childhood adiposity only effects OSA via adult adiposity. These results highlight interventions that focus on preventing and maintaining healthy adulthood body size to improve sleep helath.

## List of abbreviations

OSA: Obstructive sleep apnea
MR: Mendelian randomization
SNP: Single nucleotide polymorphism
GWAS: Genome-wide association study
BMI: Body mass index
MVP: Million Veteran Program
ICD: International Classification of Disease
LD: Linkage Disequilibrium
IVW: Inverse variance weighted
OR: Odds ratio
CI: Confidence interval

## Data Availability

GWAS Summary statistic data of childhood and adulthood body size are available through request to UK Biobank.
GWAS summary statistic of OSA can be accessed through application at: https://www.ncbi.nlm.nih.gov/gap/ under the MVP accession (phs001672).
GWAS summary statistic of Insomnia and Chronotype are requested from 23andME.

## Acknowledgements

We thank the participants of the UK Biobank, Million Veteran Program, and 23andMe study and the genome-wide association study consortia who made their summary statistics publicly available for this study.

## Author contributions

S. P. and B. B. conceived and design the study. S. P. analysed the data and wrote first draft of the manuscript. All authors took part in the interpretation and revision of the manuscript. S. P. and B. B. are accountable for the accuracy and integrity of all parts of the works.

## Ethical standards

We used publicly available summary statistics from published studies. Ethical approval and participant consent for each study is detailed in the respective publications.

## Data availability

R script for two-sample MR can be accessed through Github https://github.com/hunt-genes/mvmr_bmi_sleep.

GWAS Summary statistic data of childhood and adulthood body size are available in the ‘Additional file: Sheet 1-4 ’.

GWAS summary statistic of OSA can be accessed through application at: https://www.ncbi.nlm.nih.gov/gap/ under the MVP accession (phs001672).

GWAS summary statistic of Insomnia and Chronotype are requested from 23andME.

