## Additional file 1 for "Investigating adiposity in childhood and adulthood on later life sleep health: a lifecourse Mendelian randomization study": Additional file 1.docx

**Table S1.** Univariable and multivariable inverse variance weighted (IVW) estimates of childhood and adulthood adiposity on insomnia and morning chronotype.

| **Exposures** | **Outcomes** | **Model 1* (Univariable MR)** | | | | **Model 2* (Multivariable MR)** | | | | **Model 3* (Multivariable MR)** | | | |
| --- | --- | --- | --- | --- | --- | --- | --- | --- | --- | --- | --- | --- | --- |
|  |  | **SNPs** | **OR** | **95% CI** | **P-value** | **SNPs** | **OR** | **95% CI** | **P-value** | **SNPs** | **OR** | **95% CI** | **P-value** |
| Childhood adiposity | Insomnia | 192 | 0.94 | 0.87 to 1.03 | 0.171 | 700 | 0.70 | 0.64 to 0.77 | 6.31×10^-13^ | 692 | 0.70 | 0.64 to 0.77 | 9.21×10^-13^ |
|  | Morning chronotype | 288 | 1.16 | 1.01 to 1.33 | 0.032 | 612 | 1.36 | 1.15 to 1.62 | 4.79×10^-4^ | 616 | 1.38 | 1.16 to 1.64 | 2.75×10^-4^ |
| Adulthood adiposity | Insomnia | 420 | 1.35 | 1.25 to 1.45 | 2.67×10^-14^ | 700 | 1.58 | 1.46 to 1.72 | 2.61×10^-26^ | 692 | 1.61 | 1.47 to 1.77 | 1.82×10^-22^ |
|  | Morning chronotype | 421 | 0.87 | 0.77 to 0.98 | 0.020 | 612 | 0.80 | 0.68 to 0.92 | 0.003 | 616 | 0.73 | 0.61 to 0.87 | 3.04×10^-4^ |

Table illustrating the total (univariable MR) and direct (multivariable MR) causal estimates of childhood and adulthood adiposity on later life insomnia and morning chronotype.

*Model 1- unadjusted, Model 2- adjusted for childhood/adulthood adiposity, Model 3 – adjusted for childhood/adulthood adiposity and obstructive sleep apnea

Abbreviations: MR, Mendelian Randomization; CI, confidence interval; SNP, Single nucleotide polymorphism; OR, Odds ratio.

**Table S2.** Univariable and multivariable MR Egger estimates of childhood and adulthood adiposity on insomnia and morning chronotype.

| **Exposures** | **Outcomes** | **Univariable MR** | | | | **Multivariable MR** | | | |
| --- | --- | --- | --- | --- | --- | --- | --- | --- | --- |
|  |  | **SNPs** | **OR** | **95% CI** | **P-value** | **SNPs** | **OR** | **95% CI** | **P-value** |
| Childhood adiposity | Insomnia | 192 | 0.99 | 0.82 to 1.19 | 0.886 | 700 | 0.72 | 0.65 to 0.80 | 3.17×10^-09^ |
|  | Morning chronotype | 228 | 1.07 | 0.80 | 0.650 | 612 | 1.45 | 1.19 to 1.76 | 2.81×10^-4^ |
| Adulthood adiposity | Insomnia | 420 | 1.22 | 0.98 to 1.52 | 0.075 | 700 | 1.61 | 1.47 to 1.77 | 1.08×10^-23^ |
|  | Morning chronotype | 421 | 1.22 | 0.87 to 1.71 | 0.251 | 612 | 0.83 | 0.70 to 0.98 | 0.026 |

Abbreviations: MR, Mendelian Randomization; CI, confidence interval; SNP, Single nucleotide polymorphism; OR, Odds ratio.

**Table S3.** Univariable and multivariable MR Egger intercepts of childhood and adulthood adiposity on insomnia and morning chronotype.

| **Exposures** | **Outcomes** | **Univariable MR** | | | **Multivariable MR** | | |
| --- | --- | --- | --- | --- | --- | --- | --- |
|  |  | **SNPs** | **intercept** | **P-value** | **SNPs** | **intercept** | **P-value** |
| Childhood adiposity | Insomnia | 192 | -0.0006 | 0.605 | 700 | 0.000 | 0.364 |
|  | Morning chronotype | 228 | 0.001 | 0.539 | 612 | -0.001 | 0.224 |
| Adulthood adiposity | Insomnia | 420 | 0.001 | 0.345 | 700 | 0.000 | 0.364 |
|  | Morning chronotype | 421 | -0.004 | 0.036 | 612 | -0.001 | 0.224 |

Abbreviations: MR, Mendelian Randomization; SNP, Single nucleotide polymorphism.

**Table S4.** Univariable weighted mode and weighted median of childhood and adulthood adiposity on insomnia and morning chronotype.

| **Exposures** | **Outcomes** | **Univariable weight median** | | | | **Univariable weighted mode** | | | |
| --- | --- | --- | --- | --- | --- | --- | --- | --- | --- |
|  |  | **SNPs** | **OR** | **95% CI** | **P-value** | **SNPs** | **OR** | **95% CI** | **P-value** |
| Childhood adiposity | Insomnia | 192 | 0.97 | 0.91 to 1.03 | 0.318 | 192 | 1.04 | 0.95 to 1.13 | 0.378 |
|  | Morning chronotype | 228 | 1.12 | 0.98 to 1.29 | 0.105 | 228 | 0.74 | 0.44 to 1.22 | 0.235 |
| Adulthood adiposity | Insomnia | 420 | 1.19 | 1.13 to 0.23 | 5.16×10^-08^ | 420 | 1.10 | 1.00 to 1.21 | 0.005 |
|  | Morning chronotype | 421 | 0.85 | 0.76 to 0.95 | 0.003 | 421 | 0.84 | 0.59 to 1.21 | 0.362 |

Abbreviations: CI, confidence interval; SNP, Single nucleotide polymorphism; OR, Odds ratio.

**Table S5.** MR-Robust Adjusted Profile Score (MR-RAPS) estimates for from childhood and adulthood adiposity to insomnia and morning chronotype.

| **Exposure** | **Outcome** | **SNPs** | **OR** | **P-value** |
| --- | --- | --- | --- | --- |
| Childhood adiposity | Insomnia | 267 | 0.95 | 0.169 |
|  | Morning chronotype | 67 | 0.99 | 0.948 |
| Adulthood adiposity | Insomnia | 485 | 1.27 | 8.41×10^-12^ |
|  | Morning chronotype | 149 | 0.99 | 0.915 |

Abbreviations: MR, Mendelian randomization; SNP, Single nucleotide polymorphism; OR, Odds ratio

**Supplement Figure S1.** Leave-one-out anysis plot for Inverse Variance Weighted (IVW) estimates of childhood and adulthood adiposity on Insomnia and Morning chronotype.

A)
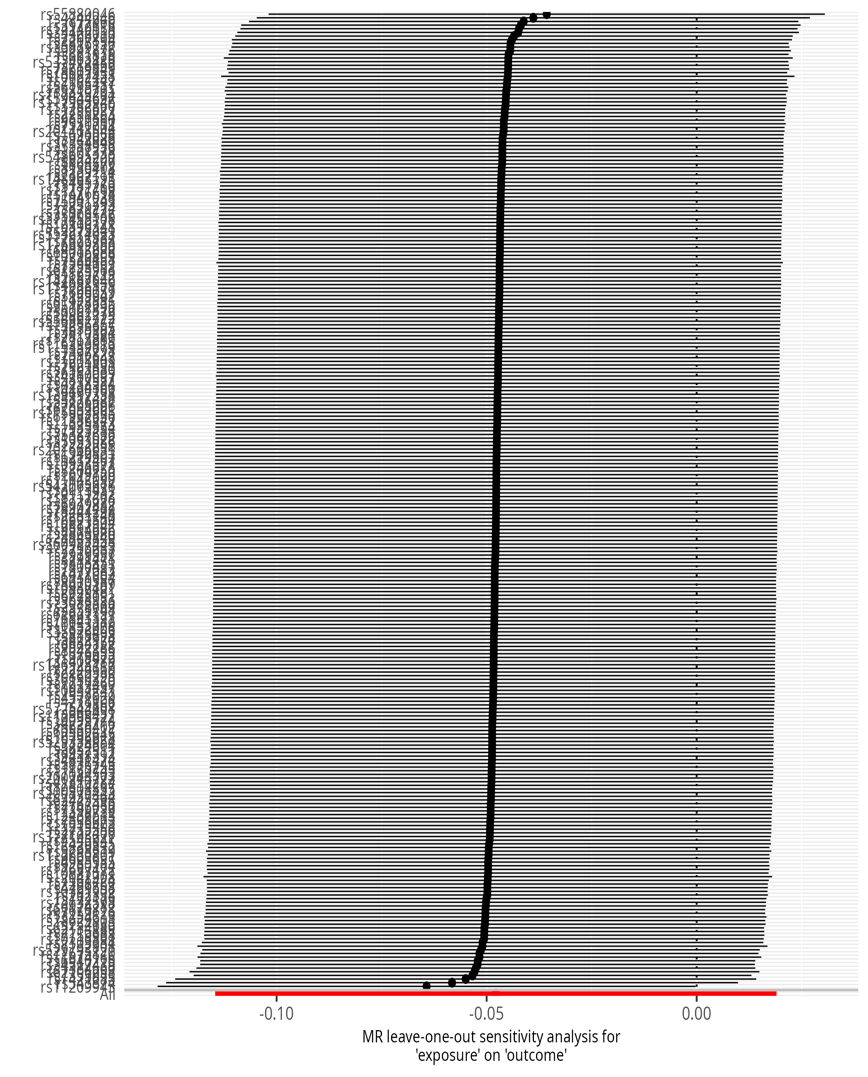
Childhood Adiposity on Insomnia	B) Adulthood Adiposity on Insomnia

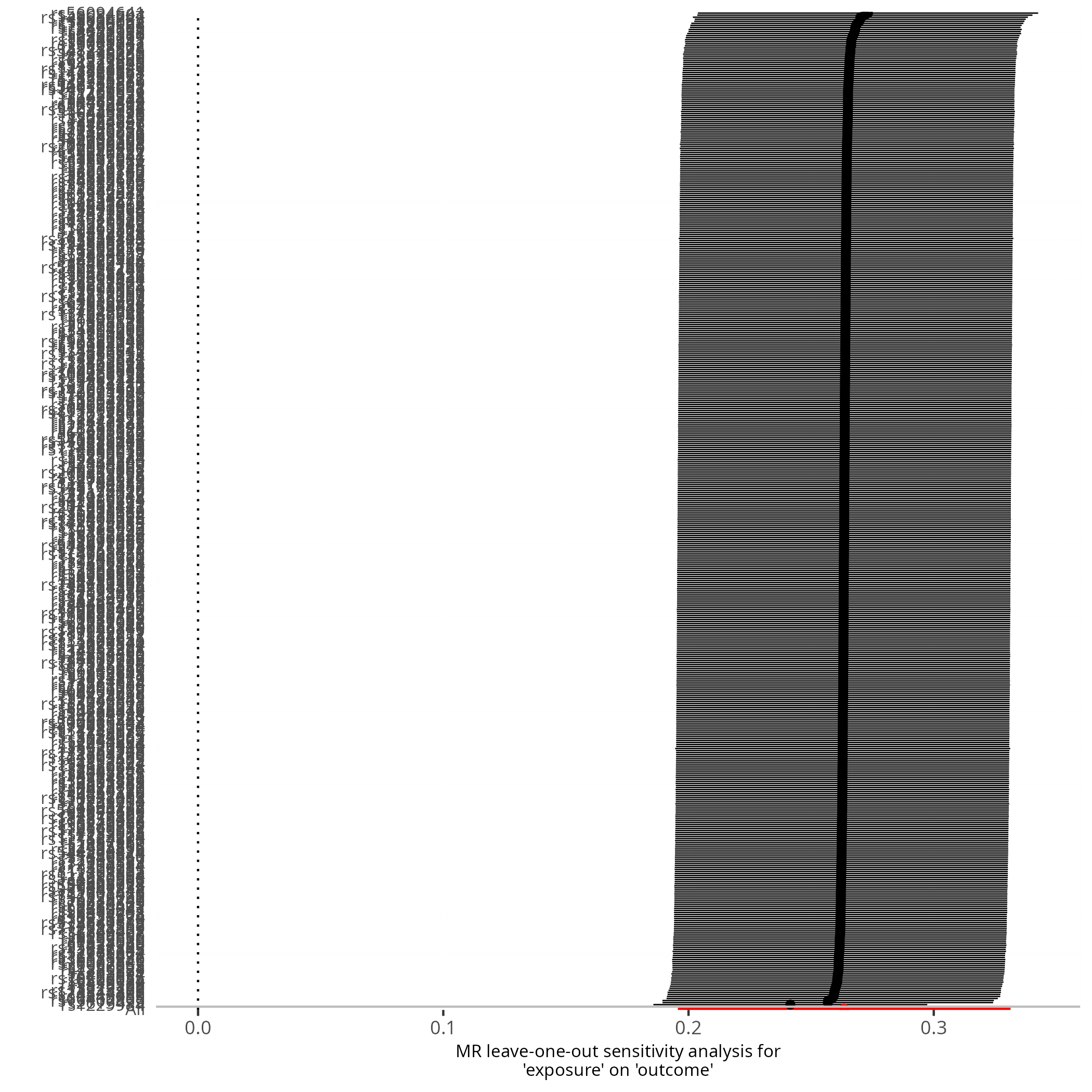

C) Childhood Adiposity on Morning chronotype D) Adulthood Adiposity on Morning chronotype

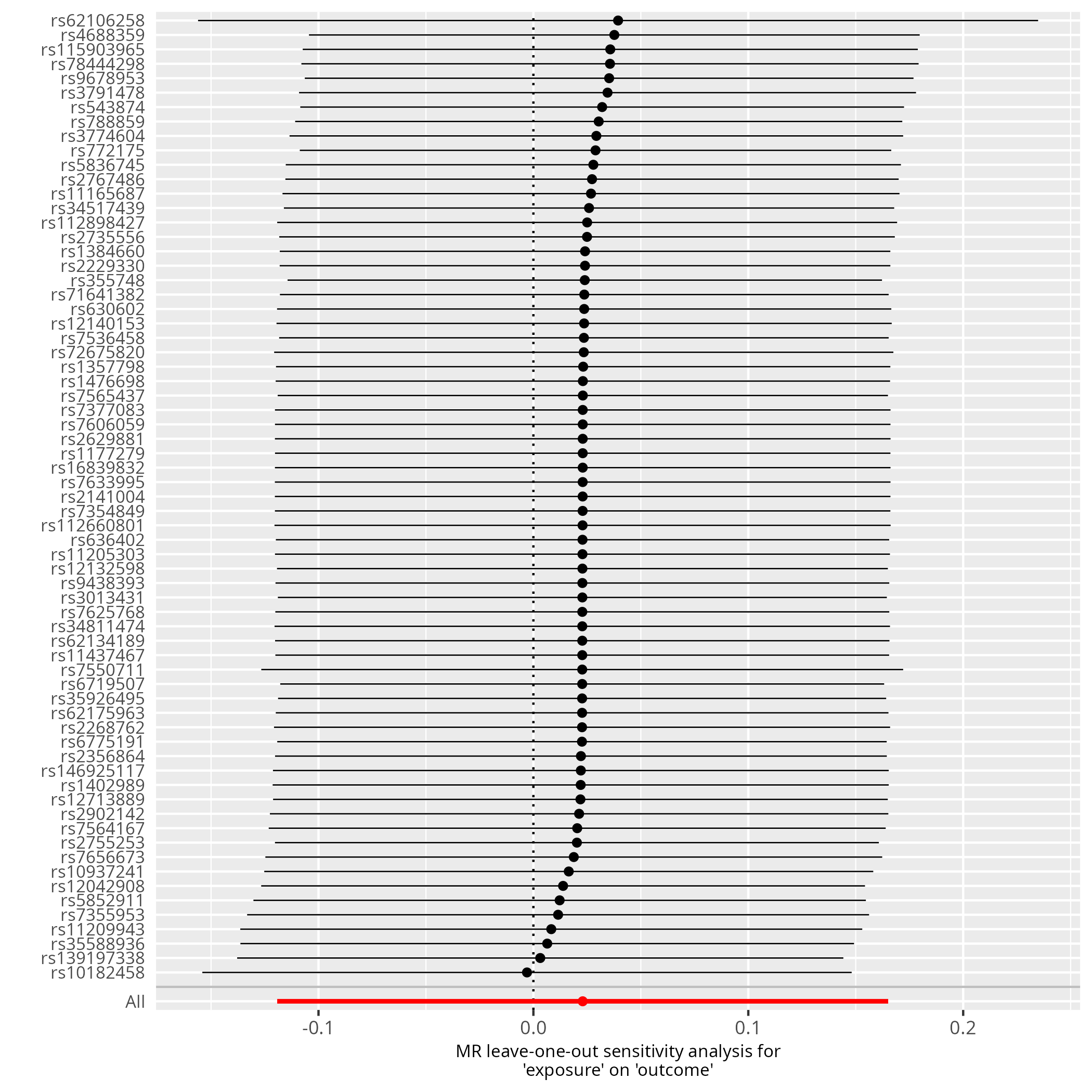

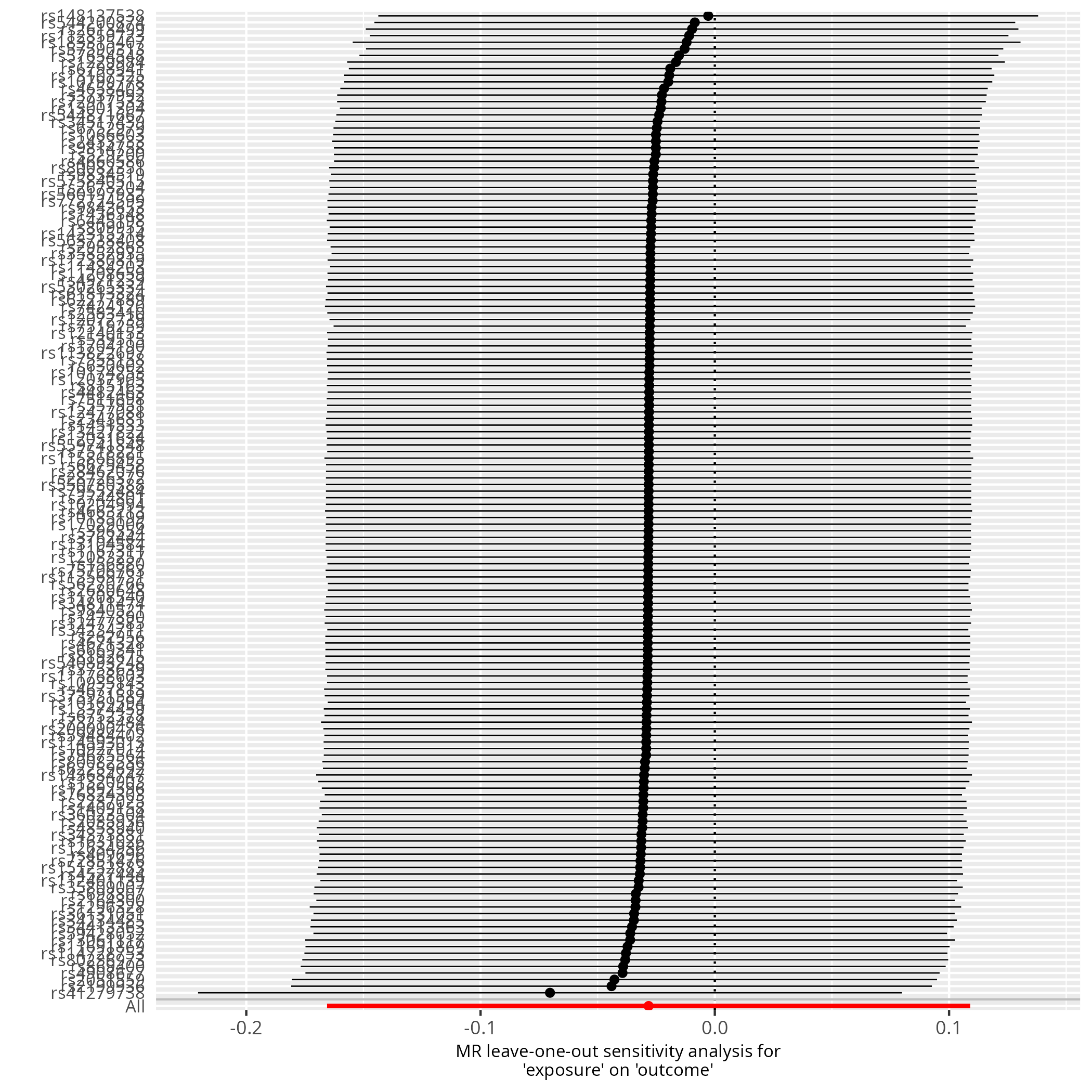

**Table S6.** Directionality test from childhood and adulthood adiposity to Insomnia and Morning chronotype.

| **Exposure** | **Outcome** | **snp_r2.exposure** | **snp_r2.outcome** | **correct_causal_direction** | **steiger_pval** |
| --- | --- | --- | --- | --- | --- |
| Childhood adiposity | Insomnia | 0.042 | 0.001 | True | <0.005 |
|  | Morning chronotype | 0.013 | 0.0003 | True | <0.005 |
| Adulthood adiposity | Insomnia | 0.063 | 0.002 | True | <0.005 |
|  | Morning chronotype | 0.020 | 0.001 | True | <0.005 |

**Table S7**. Conditional F statistics of childhood and adulthood adiposity in univariable and multivariable MR

| **Exposure** | **Outcome** | **Univariable MR** | **Multivariable MR** |
| --- | --- | --- | --- |
| Childhood adiposity | Insomnia | 65 | 13 |
|  | Morning chronotype | 67 | 14 |
| Adulthood adiposity | Insomnia | 51 | 15 |
|  | Morning chronotype | 52 | 16 |

**Table S8**. Heterogeneity test of univariable and multivariable MR

| **Exposure** | **Outcome** | **Univariable MR** | **Multivariable MR** |
| --- | --- | --- | --- |
|  |  | **Q (P-value)** | **Q (P-value)** |
| Childhood adiposity | Insomnia | 1356 (1.57×10^-174^) | 5510 (1.26×10−880) |
|  | Morningness | 896 (3.32×10^-80^) | 2396 (2.12×10^-210^) |
| Adulthood adiposity | Insomnia | 4064 (1.41×10^-13^) | 5510 (1.26×10−880) |
|  | Morningness | 1800 (8.71×10^-170^) | 2396 (2.12×10^-210^) |

**Table S9**. Univariable and multivariable Inverse Variance Weighted (IVW) estimates of Obstructive sleep apnoea on insomnia and morning chronotype.

| **Exposures** | **Outcomes** | **Univariable MR** | | | | **Multivariable MR (accounting for childhood and adulthood body size)** | | | |
| --- | --- | --- | --- | --- | --- | --- | --- | --- | --- |
|  |  | **SNPs** | **OR** | **95% CI** | **P-value** | **SNPs** | **OR** | **95% CI** | **P-value** |
| Obstructive sleep apnoea | Insomnia | 2 | 1.61 | 1.44 to 1.81 | 2.42×10^-16^ | 692 | 0.97 | 0.91 to 1.03 | 0.268 |
|  | Morning chronotype | 7 | 1.31 | 1.09 to 1.59 | 0.005 | 616 | 1.15 | 1.03 to 1.27 | 0.009 |

Abbreviations: MR, Mendelian Randomization; CI, confidence interval; SNP, Single nucleotide polymorphism; OR, Odds ratio.

**Table S10.** Univariable and multivariable Inverse Variance Weighted (IVW) estimates of childhood and adulthood adiposity on Obstructive sleep apnoea.

| **Exposures** | **Outcomes** | **Univariable MR** | | | | **Multivariable MR** | | | |
| --- | --- | --- | --- | --- | --- | --- | --- | --- | --- |
|  |  | **SNPs** | **OR** | **95% CI** | **P-value** | **SNPs** | **OR** | **95% CI** | **P-value** |
| Childhood adiposity | Obstructive sleep apnoea | 248 | 1.73 | 1.55 to 1.92 | 2.11×10^-23^ | 605 | 1.07 | 0.95 to 1.21 | 0.276 |
| Adulthood adiposity |  | 466 | 2.31 | 2.13 to 2.50 | 3.30×10^-90^ | 605 | 2.19 | 1.96 to 2.43 | 5.18×10^-41^ |

Abbreviations: MR, Mendelian Randomization; CI, confidence interval; SNP, Single nucleotide polymorphism; OR, Odds ratio.

**Table S11.** Directionality test from Obstructive Sleep Apnea to Insomnia.

| snp_r2.exposure | snp_r2.outcome | correct_causal_direction | steiger_pval |
| --- | --- | --- | --- |
| 0.0005 | 0.0000018 | True | 3.49×10^-34^ |

**Table S12.** Univariable and multivariable Inverse Variance Weighted (IVW) estimates of childhood and adulthood adiposity on sleep duration, daytime sleepiness, and daytime napping.

| **Exposures** | **Outcomes** | **Univariable MR** | | | | **Multivariable MR** | | | |
| --- | --- | --- | --- | --- | --- | --- | --- | --- | --- |
|  |  | **SNPs** | **Beta** | **95% CI** | **P-value** | **SNPs** | **Beta** | **95% CI** | **P-value** |
| Childhood adiposity | Sleep duration | 237 | -0.06 | -0.10 to -0.25 | 0.001 | 560 | 0.02 | -0.08 to 0.04 | 0.616 |
|  | Daytime sleepiness | 237 | 0.02 | 0.001 to 0.04 | 0.03 | 560 | -0.02 | -0.05 to 0.003 | 0.09 |
|  | Daytime napping | 305 | -0.04 | -0.06 to -0.01 | 0.003 | 727 | -0.13 | -0.16 to -0.10 | 7.44×10^-16^ |
| Adulthood adiposity | Sleep duration | 425 | -0.09 | -0.13 to -0.05 | 9.26×10^-06^ | 560 | -0.08 | -0.13 to -0.03 | 0.003 |
|  | Daytime sleepiness | 425 | 0.05 | 0.04 to 0.07 | 1.20×10^-11^ | 560 | 0.06 | 0.04 to 0.08 | 1.10×10^-09^ |
|  | Daytime napping | 563 | 0.08 | 0.06 to 0.09 | 1.53×10^-14^ | 727 | 0.14 | 0.12 to 0.17 | 2.15×10^-24^ |

Abbreviations: MR, Mendelian Randomization; CI, confidence interval; SNP, Single nucleotide polymorphism.

**Table S13. Questions used to define cases and control of insomnia**

| **No.** | **Cases** | **Control** |
| --- | --- | --- |
| **1.** | Have you ever been diagnosed with, or treated for, insomnia?. | Have you ever been diagnosed with, or treated for, any of the following conditions?’ (Insomnia; Narcolepsy; Sleep apnea; Restless leg syndrome) |
| **2.** | Were you diagnosed with insomnia?. | In the past 12 months, have you been newly diagnosed with any of the following conditions by a medical professional?’ (Insomnia; Sleep apnea; Migraines) |
| **3.** | Have you ever been diagnosed by a doctor with any of the following neurological conditions? (Sleep disturbance). | Have you ever been diagnosed with or treated for any of the following conditions?’ (Posttraumatic stress disorder; Autism; Asperger’s; Sleep disorder) |
| **4.** | Do you routinely have trouble getting to sleep at night?. | Have you ever been diagnosed with or treated for a sleep disorder? |
| **5.** | What sleep disorders have you been diagnosed with? Please select all that apply. (Insomnia, trouble falling or staying asleep). | Have you ever been diagnosed with or treated for any of the following conditions? (A sleep disorder) |
| **7.** | Have you ever taken these medications? (Prescription sleep aids). |  |
| **8.** | In the last 2 years, have you taken any of these medications?’ (Prescription sleep aids). |  |
